# Robust Machine Learning predicts COVID-19 Disease Severity based on Single-cell RNA-seq from multiple hospitals

**DOI:** 10.1101/2022.10.21.22280983

**Authors:** Amina Lemsara, Adrian Chan, Dominik Wolff, Michael Marschollek, Yang Li, Christoph Dieterich

## Abstract

Coronavirus disease 2019 (COVID-19) has a highly variable disease severity. Possible associations between peripheral blood signatures and disease severity have been investigated since the emergence of the pandemic. Although several signatures were identified based on exploratory analyses of single-cell omics data, there are no state-of-the-art validated models to predict COVID-19 severity from comprehensive transcriptome profiling of Peripheral Blood Mononuclear Cells (PBMCs). In this paper, we present a computational workflow based on a Multilayer perceptron network that predicts the necessity of mechanical ventilation from PBMCs single-cell RNA-seq data. The study includes patient cohorts from Bonn, Berlin, Stanford, and three Korean medical centers. Training and model validation are performed using Berlin and Bonn samples, while testing is performed on completely unseen samples from the Stanford and Korean datasets. Our model shows a high area under the receiver operating characteristic (AUROC) curve (Korea: 1 (CI:1-1), Stanford: 0.86 (CI:0.81-0.9)), proving our model’s robustness. Moreover, we explain our model’s performance by identifying gene loci and cell types, which are most critical for the classification task. In summary, we could show that the expression of 15 genes and the cell type proportion of 29 PBMC classes distinguish between COVID-19 disease states.

**Graphical Abstract:** 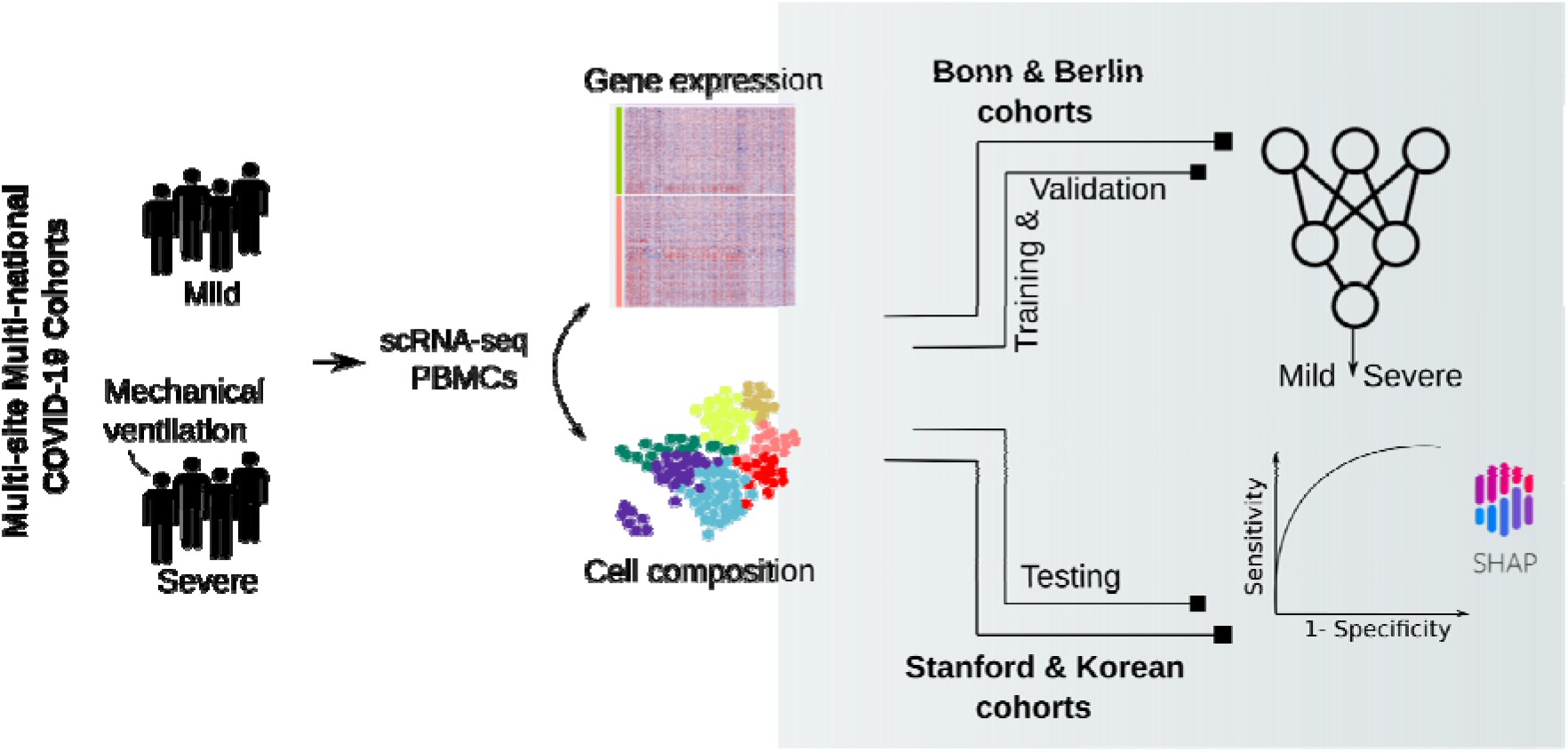

## INTRODUCTION

Characterizing COVID-19 severity from genomic expression data is important to uncover the underlying biological mechanism of the disease and to identify the optimal treatment for each patient. Overall, the acuity of COVID-19 infection varies broadly among individuals, ranging from asymptomatic to fatal. In some cases, the disease can cause a life-threatening condition characterized by systemic inflammation accompanied by acute respiratory distress syndrome and multi-organ failure (Chen et al., 2020; Wu et al., 2020). Such cases need immediate intervention and intensive care. It is thus essential to have a diagnostic tool that will enable an assessment of the disease state, independent of clinical observations.

The biological basis of severe COVID-19 symptoms is currently not fully understood. Identifying molecular disease signatures underlying the clinical manifestations is a crucial step in designing therapeutic strategies. Previous studies have highlighted the critical role of immune and inflammatory responses (García, 2020; Li et al., 2022). Systemic reviews show that severe COVID-19 cases are characterized by several abnormal laboratory markers (Gupta et al., 2020; Samprathi and Jayashree, 2020), including the hyperactivation of inflammatory markers in the serum such as cytokine, C-reactive protein, D-dimer, fibrinogen, and lactate dehydrogenase among others. Such observations are often accompanied by changes in cell proportions of hematologic and immune cell types, leading to lymphopenia, leukocytosis, neutrophilia, and thrombocytopenia.

The use of single-cell sequencing technology has enhanced the understanding of the biological characteristics of COVID-19 infection and has uncovered potential cellular and molecular signatures. To date, many studies addressing the contribution of peripheral blood cells to COVID-19 susceptibility and severity have been published (Consortia et al., 2020). Notably, the immunity landscape is different between severe and non-severe cases (Mukund et al., 2021; Ren et al., 2021; Stephenson et al., 2021). Single-cell studies provide evidence of the association of poor clinical outcomes with reduced lymphocyte as well as increased neutrophil counts (Hasan et al., 2021). Severe COVID-19 is characterized by the suppression and exhaustion of the T cell compartment (Diao et al., 2020; He et al., 2021; Kreutmair et al., 2021), the extrafollicular B cell activation (Woodruff et al., 2020), the increased levels of plasmablasts, and platelets (Barrett et al., 2021; Bernardes et al., 2020), as well as the reduced function of natural killer cells (NK) (Björkström and Ponzetta, 2021) and dendritic cells (DC) (Kvedaraite et al., 2021). Classical monocytes (Merad and Martin, 2020) have been shown to display a type 1 IFN inflammatory signature (Lee et al., 2020) and low expression of HLA-DR (Schulte-Schrepping et al., 2020). The granulocyte compartment is also altered in severe COVID-19, displaying neutrophils with an immunosuppression signature (Schulte-Schrepping et al., 2020). All together, identifying those immune signatures underlying severe COVID-19 infections represents an important step forward to developing new and more effective ways of treating this debilitating disease.

Although various insights have been revealed from single-cell data thus far, effective integration of these data into routine clinical diagnoses and personalized medicine has been challenging (Jovic et al., 2022). The latter is primarily hindered by the high cost of single-cell experiments. The limited sample sizes of single-cell experiments presents a constraint for computational models as it reduces their robustness and reliability. Although large-scale platforms containing single-cell data have been made available to the public, computational algorithms that can effectively handle the data and predict clinical states are only emerging.

In this manuscript, we present an MLP-based framework that predicts the disease severity of COVID-19 samples using single-cell RNA sequencing (scRNA-seq) data. Our classification is binary based on the necessity of mechanical ventilation (MV): samples are either classified as “mild” (i.e., no ventilation) or “severe”. We integrate information on Cell Composition (CC) and Gene Expression (GE) to exploit two important aspects of scRNA-seq data in this classification task. Our model was trained and validated on a 77-samples dataset and tested on two external datasets with a total of 39 samples. It is important to note that the testing data are completely independent from the training process, which demonstrates the model’s generalizability across different sites and its great potential as a tool for clinical applications. Finally, we use the trained model to extract discriminative features (cell types and marker genes) and investigate them in light of existing biological knowledge.

## RESULTS

### MLP model yields an accurate prediction of severity and generalizes to unseen Data

To predict the severity of COVID-19 cases from scRNA-seq data, we designed an MLP-based framework integrating the GE and CC information per sample. A schematic representation of the entire workflow is shown in Figure 1. In brief, our framework comprises two parts: Feature extraction and model training. The first step aims to construct two modalities of data from scRNA seq data: the expression of the top genes differentially expressed between the two conditions of interest (Mild and Severe) and the cell types’ proportion for each sample. This information serves as input to train a model, which is based on an MLP network to predict disease severity. The joint MLP-based model was trained and validated by 30 times holdout cross-validation on 77 samples from the Bonn and Berlin cohorts (Schulte-Schrepping et al., 2020). Then, its performance was tested on two unseen cohorts, respectively, from the Stanford (Wilk et al., 2021) and Korean (Lee et al., 2020) studies. In addition, joint model performance was compared to a one-modality-based model, where the MLP network considers only either GE or CC as input.

**Figure 1.**
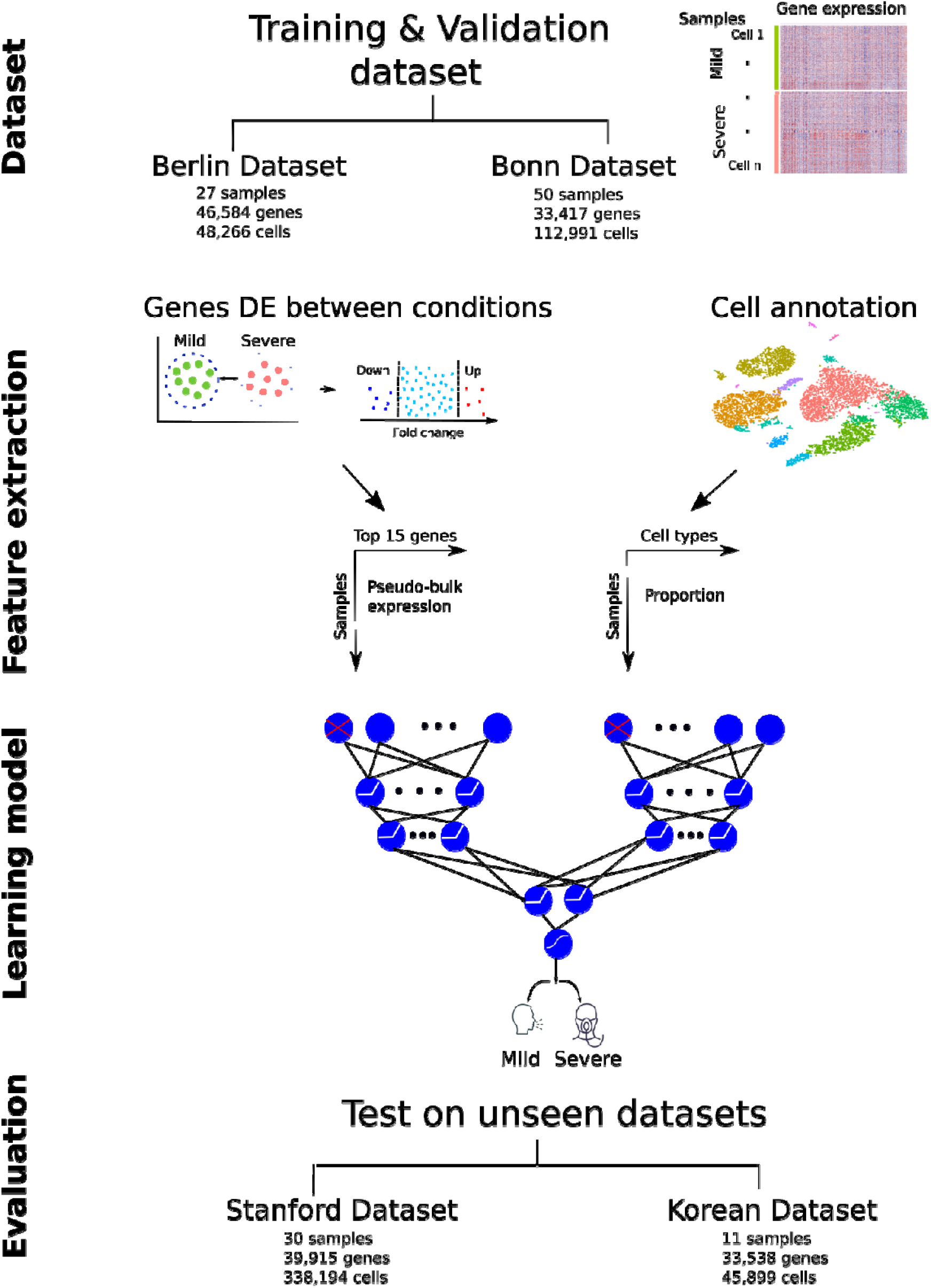
Schematic representation of the prediction workflow. Dataset: scRNA-seq data of 77 samples were collected from Bonn and Berlin cohorts and fed as input for the next step. Model construction: two matrices representing the expression of 15 genes and the proportion of 29 cell types per sample were built from the scRNA seq input data and provided as input for an MLP model to predict severity. Evaluation: the MLP-based model was evaluated on the validation set and two scRNA-seq datasets from Stanford and Korean cohorts. Further details about datasets can be found in tables: S1 and S2. A detailed architecture of the MLP model can be found in figure S1.

The Bonn cohort comprises gene expression data of 50 patient samples (25 mild and 25 severe cases), with an average of 2,260 cells per sample, whereas the Berlin cohort has 27 samples (13 Mild and 14 Severe), with an average of 1,788 cells per sample. 80% of this combined set is used to train our MLP-based severity prediction model. Then, the model is evaluated using samples from the Stanford (14 Mild and 14 Severe) and Korean (5 Mild and 6 Severe) cohorts, with an average of 11,273 and 4,172 cells per sample respectively. All samples are accompanied by information on MV necessity assigned at the collection time point. Based on the MV, we stratify the patients’ disease acuity into two classes: Mild (WHO score <=4, non-ventilated) and Severe (WHO score >4, ventilated). Further details on data characteristics can be found in Table S1 and Table S2.

We transferred cell type annotations from a large, publicly available dataset of human PBMCS (Hao et al., 2021), representing about 260,000 cells using the Seurat reference mapping (see Methods section). This allows for better comparison across datasets, fast annotation, and reproducibility. As a result, we annotate up to 31 cell types within each dataset, including T cells (33%-43%) and Monocytes (30%-37%) as the major cell types (Figure 2, Table S3). Based on the cell annotation, we computed the proportion of each cell type per sample. We excluded Doublet and Eryth types from subsequent analyses. In addition to the cell proportion, we compute the gene expression average, the so-called “pseudo bulk” profile, by aggregating cell-level counts into sample-level counts (Figure 1). Due to the sample size limit characterizing single-cell data, we opted for feature (genes) selection to reduce the complexity of the model. Herein, we are interested particularly in the top genes differentially expressed between conditions. Based on the Area Under the Receiver Operating Characteristics (AUROC), area under the precision-recall curve (AUPRC) and Accuracy, we evaluated the prediction model using various numbers of top genes; accordingly, with 15 top genes, the model shows the best results on the validation set (Figure S1).

**Figure 2.**
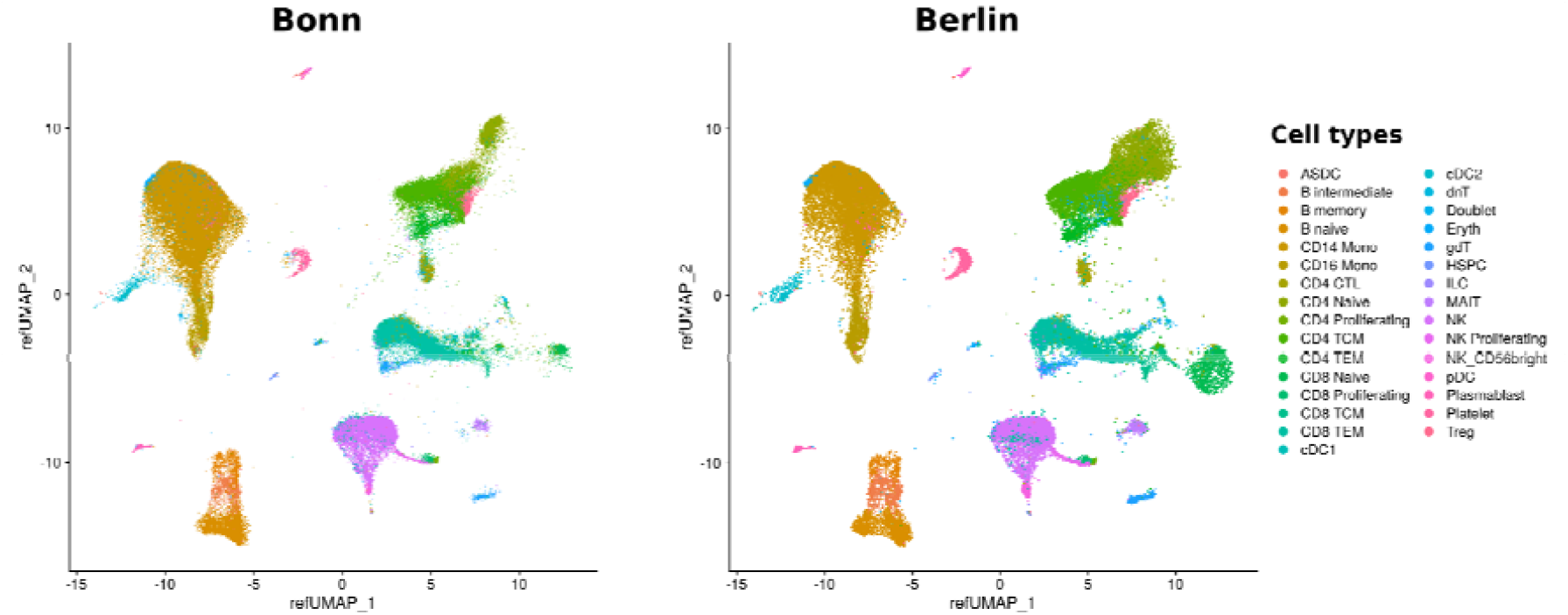
Cell annotation of Bonn and Berlin datasets based on Seurat reference mapping represented by UMAP. Doublets and Erythrocytes are excluded from further analyses. Cell composition for all datasets can be found in Table S3.

In summary, we construct two input data matrices (modalities) representing the gene expression (77 samples x 15 genes) and the cell proportion (77 samples x 29 cell types) across samples. We randomly assign 80% of the samples for the training and 20% for the validation. We design an MLP network of 4 hidden layers to estimate the severity probability. Model architecture is shown in Figure S1 and described in the Methods section. Furthermore, we employ the dropout (Srivastava et al., 2014) and early-stopping (Prechelt, 2012) techniques during the training process to control overfitting (see Methods section for further details). The resulting model allows the classification of samples into mild and severe conditions with a decision cut-off of 0.5.

We evaluated the model using several metrics. Figure 3 shows performances with a 95% confidence interval (CI). Overall, we obtain high AUROC, AUPRC, and accuracy across 30 random samplings (Table 1).

**Table 1:** Performance statistics of the joint MLP-based model on the validation set, Korean and Stanford datasets in terms of the Area Under the Receiver Operating Characteristics (AUROC), Area Under the Precision-Recall Curve (AUPRC), and Accuracy. Thirty sampling replicates were used to assess the prediction performance with 95% CI. Further results can be found in the supplementary material (Table S4).

|  | AUROC | AUPRC | Accuracy |
| --- | --- | --- | --- |
| Validation set | 1.0 (0.99-1.0) | 1 (0.99-1.0) | 0.96 (0.94-0.99) |
| Korean dataset | 1.0 (1.0-1.0) | 1.0 (1.0-1.0) | 0.9 (0.8 -0.9) |
| Stanford dataset | 0.86 (0.8-0.89) | 0.89 (0.84-0.92) | 0.77 (0.70-0.86) |

**Figure 3:**
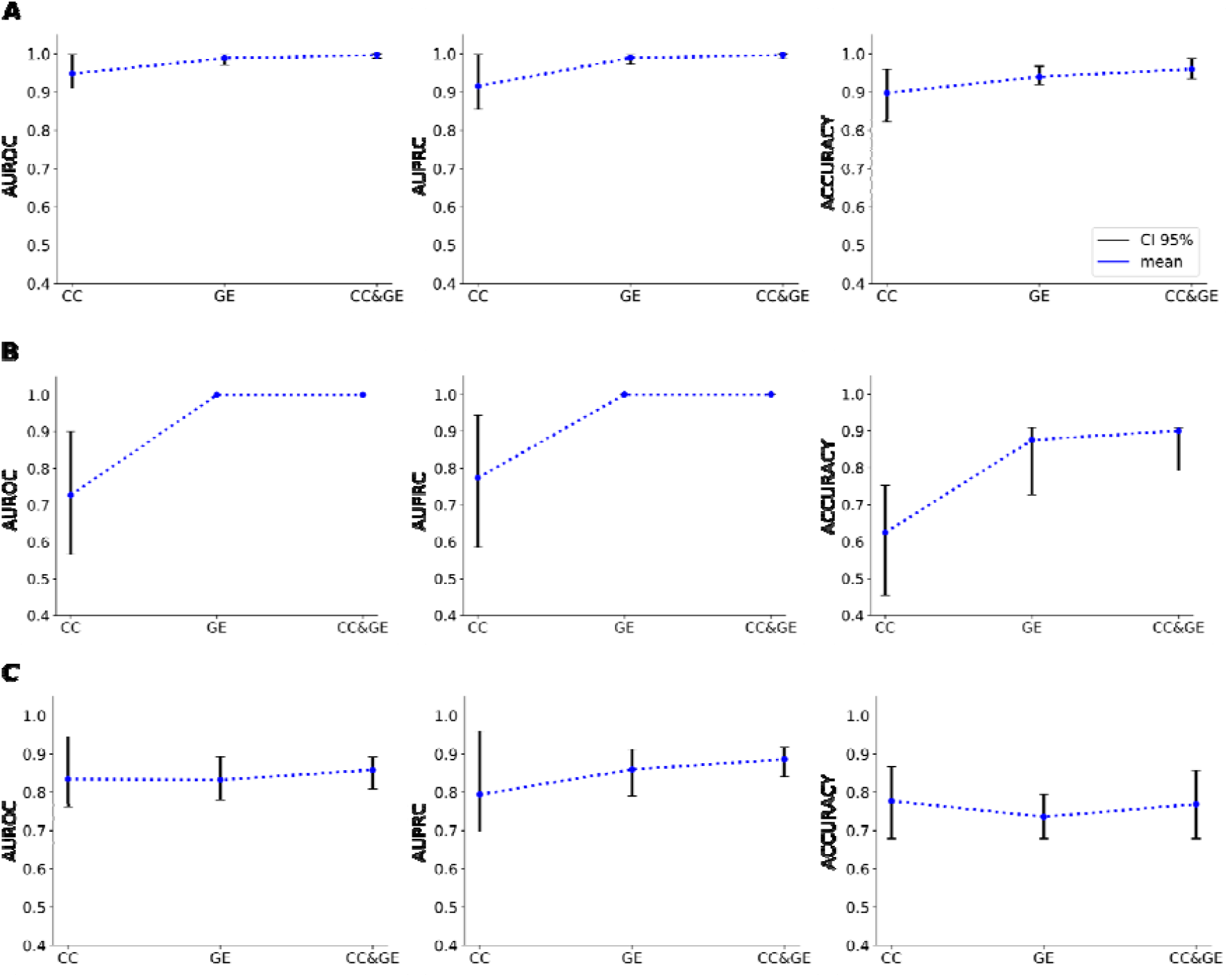
Performance of the MLP-based model on the validation set (A) Korean (B) and Stanford (C) datasets. Three models were evaluated: Cell composition-based mode (CC), Gene expression-based model (GE), and joint modal (CC&GE) using the Area Under the Receiver Operating Characteristics (AUROC), Area Under the Precision-Recall Curve (AUPRC), and Accuracy. Thirty sampling replicates were used to assess the prediction performance with 95% CI. Further results can be found in the supplementary material (Table S4).

To demonstrate the importance of combining both modalities (CC and GE) as input features, we compared the prediction performance of the joint MLP-based model against each individual modality, separately (Figure 3 and Table S4). Indeed, compared to the prediction based on one modality of features, the joint model achieves better AUROC, AUPRC, and accuracy, indicating that our method can successfully and efficiently capture information from scRNA-seq data.

Overall, our results show that the model can assess the disease state of COVID-19 patients and is generalizable to completely unseen data collected at completely different sites, which suggests that the model captures the key markers (patterns) of severity.

### MLP-based prediction allows for interpretable classification

We computed SHAP values (Lundberg and Lee, 2017; Lundberg et al., 2018) to understand better the relevance of individual features (Cell types and Genes) for disease severity prediction. In Figure 4, we show bar plots representing the top 15 features from both modalities ranked according to the average across the 30 samplings of the absolute mean of the SHAP values. For example, the top-ranked RETN gene discriminates between severe and mild conditions. Resistin (RETN) is known to correlate positively with poor prognosis in COVID-19 patients and is an important predictor of mechanical ventilation necessity (Perpiñan et al., 2022). It is also known to play a crucial role in neutrophil activation and is associated with bexcessive proinflammatory activation (Jiang et al., 2014). RETN has also been identified together with lipocalin-2 (LCN2), the second top predictor for Korean samples, as the most important factors in distinguishing critical illness in COVID-19 in a previous machine learning-based finding ((Meizlish et al., 2021). In addition, the model highlights the importance of the platelet marker gene PPBP, which was reported to be a strong negative predictor of mechanical ventilation in hospitalized patients ((Yatim et al., 2021). Interestingly, we observed the high contribution of Platelet, Natural Killer (NK) cells, and T cell types, known to be involved in disease progression ((Barrett et al., 2021; Björkström and Ponzetta, 2021; Stephenson et al., 2021; Yatim et al., 2021).

**Figure 4:**
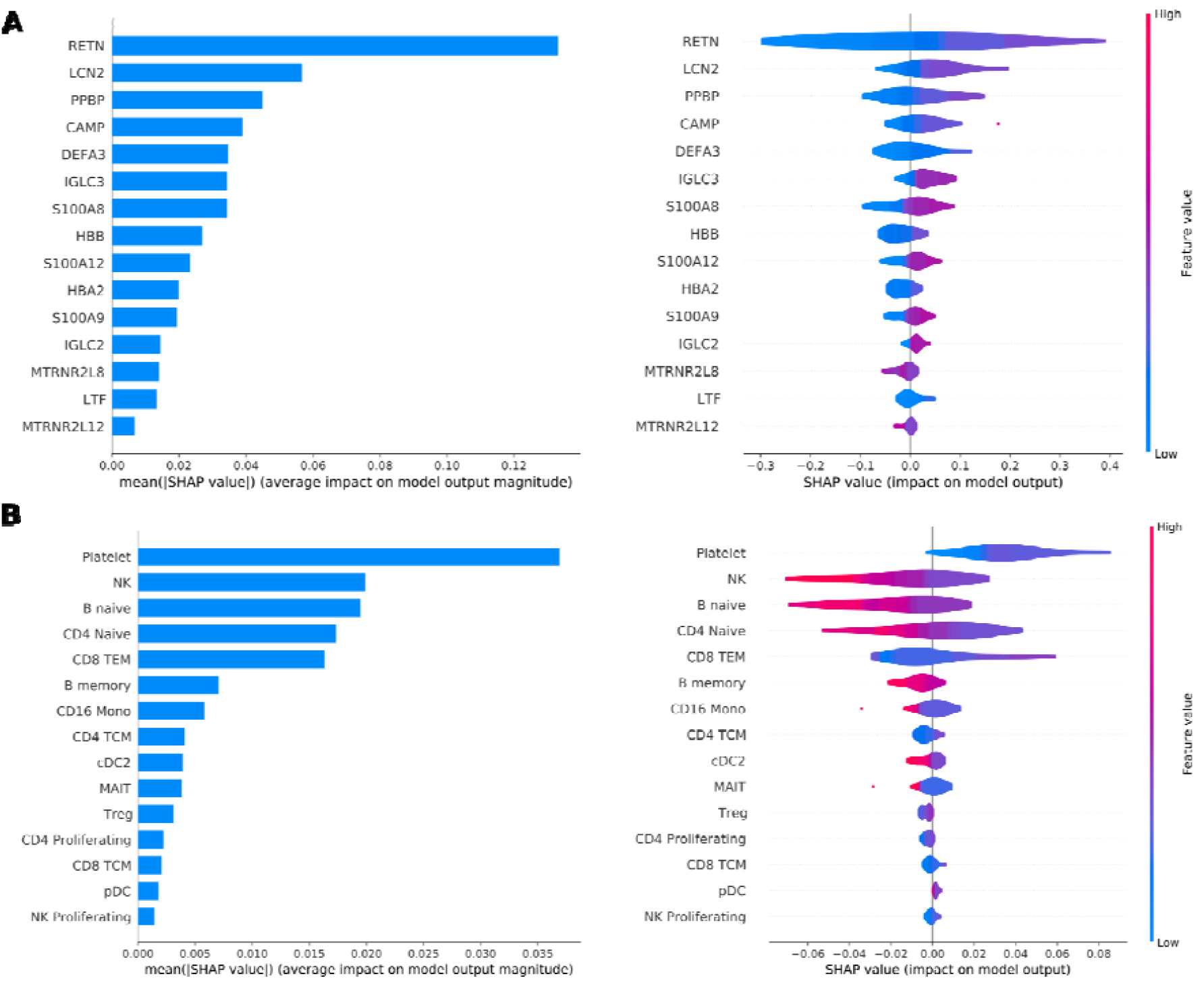
Feature importance based on SHapley Additive exPlanation (SHAP) values evaluated on the Korean dataset. Bar plots represent the average across the 30 samplings of the mean absolute SHAP values to illustrate global feature importance (left). The violin plots show the direction of the relationship between each feature and the prediction outcome (right). The color in the violin plot represents the average feature value at that position. Features with many instances in red with SHAP values greater than 0 contribute positively to the prediction, while those with many blue instances decrease the prediction. Further results can be found in the Supplementary material (Figure S2).

## DISCUSSION

Single-cell RNA sequencing enables the transcriptomic profiling of individual cells in COVID-19, leading to the characterization of heterogeneous cell populations and the identification of specific disease signatures. While scRNA-seq has been used extensively to provide insights into COVID-19 infection and its severity, it has not been used for disease prediction or diagnostics. This is mainly due to its high cost as well as the complexity and sparsity of the data. In this work, we made use of PBMC scRNA-seq data from different hospitals and constructed a robust and reliable model for predicting disease severity (current mechanical ventilation) in patient samples. We have shown that our model achieves high AUROC, AUPRC, and accuracy on independent external datasets. Notably, the MLP-based model effectively captures biological markers that lead to a better interpretability for its decisions. Collectively, the results show that our model can predict COVID-19 severity and generalizes to independent datasets. Our model (re-)discovers many aspects of previous biological knowledge in terms of marker genes and sample cell type composition. We believe that our findings contribute to the advancement of precision medicine efforts in COVID-19, specifically once molecular profiling of patient samples will be implemented in clinical care. To successfully pursue this vision, additional well-documented (at the molecular and clinical levels) samples will be necessary to train and evaluate computational models. Another future direction of our research is to integrate multi-omics data. For example, proteomics data have been employed to predict the current mechanical ventilation status based and showed high performance and generalizability (Demichev et al., 2021). While such multi-modalities data promise to provide rich insight into complex systems, integrative computational tools should reflect the biological complexity and learn from single-cell data collected across cells, experimental conditions, as well as molecular, spatial, and temporal modalities (Lance et al., 2022).

## METHOD DETAILS

The R Seurat package version 4.1.0 was used for data normalization, pseudo-bulk differential expression analysis, label transfer, and UMAP visualizations. The MLP model was developed using Keras version 2.9.0 and Tensorflow version 2.9.1.

### 1. Single-cell RNA Sequencing Datasets

We used publicly available scRNA-seq data derived from PBMCs of COVID-19 samples. Count gene expression data of Bonn and Berlin cohorts were obtained from ((Schulte-Schrepping et al., 2020). Samples were retrieved from patients recruited at the University Hospital Bonn and the Charité Universitätsmedizin of Berlin, respectively. Single-cell RNA seq raw dataset of patients enrolled in the Stanford University COVID-19 Biobanking studies and the three Korean medical centers (Asan Medical Center, Severance Hospital, and Chungbuk National University Hospital) were retrieved from the gene expression omnibus (GEO) under the accession numbers *GSE174072* and *GSE149689*, respectively. Data was already aligned to the reference genome. Bonn and Berlin cohorts were used to train and validate our prediction model, and Stanford and the Korean cohorts were used for evaluation. The linked metadata contains clinical information, including the World Health Organization (WHO) clinical ordinal scale (8: died; 7: invasive MV + additional support; 6: intubation + invasive MV; 5: non-invasive MV or high-flow nasal cannula (HFNC); 4: hospitalized with oxygen mask or nasal prongs; 3: no oxygen therapy; 2: activity limitation; 1: no activity limitation) or the necessity of MV. Note that WHO 1-4 represents samples without MV need and WHO 5-8 represents samples with MV need; thereby, we split samples into two classes based on the MV: Mild and Severe.

### 2. Quality control of scRNA-seq data

Whereas Stanford and Korean original studies provide raw data, Bonn and Berlin cohorts contain filtered data. Thus, to enable consistency, we apply the same filter parameters as proposed in ((Schulte-Schrepping et al., 2020). Cells that had > 25% mitochondrial gene reads, > 25% *HBA/HBB* gene reads, < 250 UMIs or >5,000 UMIs, and < 500 detected transcripts were considered to be of low quality and removed from further analysis.

### 3. Automated Annotation of Datasets

Seurat provides a single-cell reference mapping function to map new query datasets to a previously annotated reference (https://satijalab.org/seurat/articles/integration_mapping.html). We used this strategy to transfer cell type labels from a public annotated dataset to our training and testing datasets. First, we identified anchors between the query and reference datasets using precomputed supervised principal components on the reference dataset (*FindTransferAnchors*). Then, we transferred cell type labels to each cell of the query dataset via the previously identified anchors (*MapQuery*).

The reference is a multimodal dataset of the immune system representing about 260,000 cells represented by two modalities: scRNA-seq data and proteins with 228 antibody panels. Based on a WNN integration, they annotated cells at three levels yielding: 8 categories (level 1), 30 categories (level 2), and 57 categories (level 3), encapsulating all major and minor immune cell types. We consider level 2 as our reference to label our query datasets.

### 4. Differential Expression Analysis and Marker Genes Identification

Differential expression (DE) tests were performed using Seurat function (*FindMarkers*) with Wilcoxon Rank Sum test. Marker genes were identified by applying the DE tests for upregulated and downregulated genes between conditions (Mild & Severe).

### 5. Prediction of severity

#### Gene selection

Top-ranked genes sorted by log-fold changes were selected. Various numbers of top DE genes were tested (5,10,15,20) to evaluate the robustness of the model (figure S1); however, the model’s performances showed no improvement in AUC, AUPRC, and accuracy scores when the number of top features increased to 20. Hence, the key features were the 15 top DE genes. Then, we average their expression across the single cells to generate a pseudo-bulk expression for each sample.

#### MLP Network Architecture

We designed an MLP model to process input data from two branches (modalities) and estimate a score representing the probability of being severe (Figure S3). The input branches represent the expression of the 15 selected genes in the scale 0-to-1 and the proportion of 29 identified cell types for each sample. We used three hidden layers apart from the input layer and output layer. The first two hidden layers comprised {10,4} nodes per branch. Then, we merge the two branches to estimate the third hidden layer composed of {4} nodes and finally output a probability score using a layer of one node. To allow the modeling of non-linear patterns, we used ReLU (Rectified Linear Unit) as the activation function for the hidden nodes and Sigmoid for the output layer. Based on the output score, we classify samples into Mild (<0.5) and Severe (=>0.5).

#### Parameter Optimization

The model parameters were estimated using Adam optimizer (Kingma and Ba, 2017) and the default learning rate 0.001. The optimization process updates the model parameters for each sample in the training dataset (80%) and performs validation on the left-out dataset (20%) chosen randomly using ‘*random*.*sample’*, providing that the training subset preserves the proportion of WHO score modalities within the whole dataset. The parameters with the lowest cross-entropy define the final model used to predict the severity score of samples from Berlin and Stanford datasets. We considered the dropout of 20% of input nodes to avoid overfitting and improve robustness. We also considered the early stopping of the model training when the loss value stops improving after 100 iterations with a minimum percent improvement on the validation set error of 0.001. The best-performing model on the validation set was saved for the prediction during the learning process.

### 6. Evaluation Metrics

We assessed the ability of the model described in the previous section to predict the severity and classify samples as Mild and Severe using the Area Under the Receiver Operating Characteristics (AUROC) and Area Under the Precision-Recall Curve (AUPRC) calculated on the estimated score using *roc_auc_score average_precision_score* respectively from the scikit-learn python package. In addition, we evaluated the accuracy, precision, recall and F1-score on a 0.5 cut-off of the output score. We performed 30 samplings for all the evaluations and computed performances at a 95% confidence interval.

### 7. Feature Contribution

To evaluate the impact of individual features from both modalities (CC & GE) on the prediction score we learned for each sample, we used (Deep) SHAP values. SHAP values are a metric based on game theory that attempts to quantify the contribution of each variable to an outcome. SHAP results in a sample-specific score that may be positive or negative. The mean of the absolute SHAP values per feature is used to score the overall impact of a feature on the prediction score.

## Data Availability

We used publicly available scRNA-seq data derived from PBMCs of COVID-19 samples. Count gene expression data of Bonn and Berlin cohorts were obtained from
Schulte-Schrepping, J., Reusch, N., Paclik, D., Bassler, K., Schlickeiser, S., Zhang, B., Kraemer, B., Krammer, T., Brumhard, S., Bonaguro, L., et al. (2020). Severe COVID-19 Is Marked by a Dysregulated Myeloid Cell Compartment. Cell 182, 1419-1440.e23. https://doi.org/10.1016/j.cell.2020.08.001.
Samples were retrieved from patients recruited at the University Hospital Bonn and the Charite Universitaetsmedizin of Berlin, respectively. Single-cell RNA seq raw dataset of patients enrolled in the Stanford University COVID-19 Biobanking studies and the three Korean medical centers (Asan Medical Center, Severance Hospital, and Chungbuk National University Hospital) were retrieved from the gene expression omnibus (GEO) under the accession numbers GSE174072 and GSE149689, respectively.

https://github.com/dieterich-lab/ImmunOMICS

https://doi.org/10.5281/zenodo.6811191

## DATA AND CODE AVAILABILITY

All data files used in our workflow are available via this link: (https://doi.org/10.5281/zenodo.6811191)

A snakemake pipeline of the tool is deposited on GitHub: https://github.com/dieterich-lab/ImmunOMICS. A docker image of all dependencies has been built and deposited in DockerHub (https://hub.docker.com/repository/docker/aminale/immun2sev). Seurat mapping requires high memory usage (>100 Gb) thus, we recommend using Singularity (or Conda) designed specifically for High-Performance Computing.

## Acknowledgments

We would like to thank Harald Wilhelmi for maintaining the HP cluster of the Dieterich Lab and for his help in setting up Docker and Singularity solutions. We would like to thank Etienne Boileau, Manoj Kumar Gupta, Zhenhua Zhang, Alexandra Schimpf, Birgit Heinz, Florian Seidel, Martin Peuker, and Naima Laharnar for fruitful discussions.

## Author Contributions

Conceptualization, A.L. and C.D.; Methodology, A.L.; Formal Analysis/Software, A.L.; Writing

-Original Draft, A.L.; Writing - Review & Editing, A.L., A.C., D.W., M.M., Y.L. and C.D.; Funding Acquisition, M.M., Y.L., and C.D.; Supervision, C.D.

## Declaration of Interests

The authors declare no competing interests.

## Supplemental Information

**Figures** https://docs.google.com/document/d/19Urx5sQYJ4Q0XVbqBjxysYxkCkfvURBDHrRN6SRXtAE/edit?usp=sharing

**Tables** https://docs.google.com/spreadsheets/d/113A6LgqqJTCbKp99PAAj3xK76B1nhFoWHvDupLKeQCM/edit?usp=sharing

## Notes

### Competing Interest Statement

The authors have declared no competing interest.

### Funding Statement

This study was supported by the Netzwerk Universitaetsmedizin Germany through the project CODEX+

### Author Declarations

We used publicly available scRNA-seq data derived from PBMCs of COVID-19 samples. Count gene expression data of Bonn and Berlin cohorts were obtained from Schulte-Schrepping, J., Reusch, N., Paclik, D., Bassler, K., Schlickeiser, S., Zhang, B., Kraemer, B., Krammer, T., Brumhard, S., Bonaguro, L., et al. (2020). Severe COVID-19 Is Marked by a Dysregulated Myeloid Cell Compartment. Cell 182, 1419-1440.e23. https://doi.org/10.1016/j.cell.2020.08.001. Samples were retrieved from patients recruited at the University Hospital Bonn and the Charite Universitaetsmedizin of Berlin, respectively. Single-cell RNA seq raw dataset of patients enrolled in the Stanford University COVID-19 Biobanking studies and the three Korean medical centers (Asan Medical Center, Severance Hospital, and Chungbuk National University Hospital) were retrieved from the gene expression omnibus (GEO) under the accession numbers GSE174072 and GSE149689, respectively.

