## Supplementary Figures for "Robust Machine Learning predicts COVID-19 Disease Severity based on Single-cell RNA-seq from multiple hospitals"

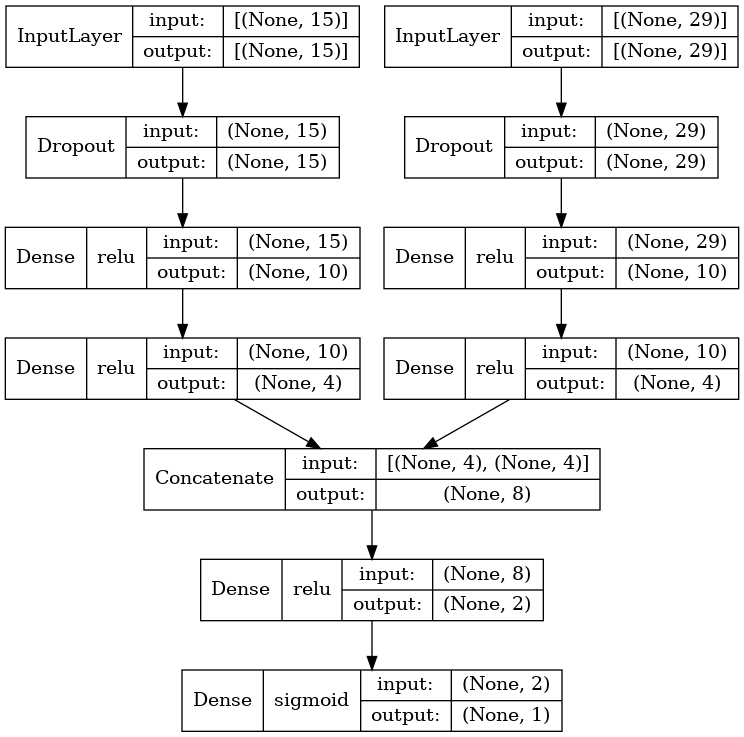

Figure S1: Architecture of the joint MLP model (Related to figure 1).

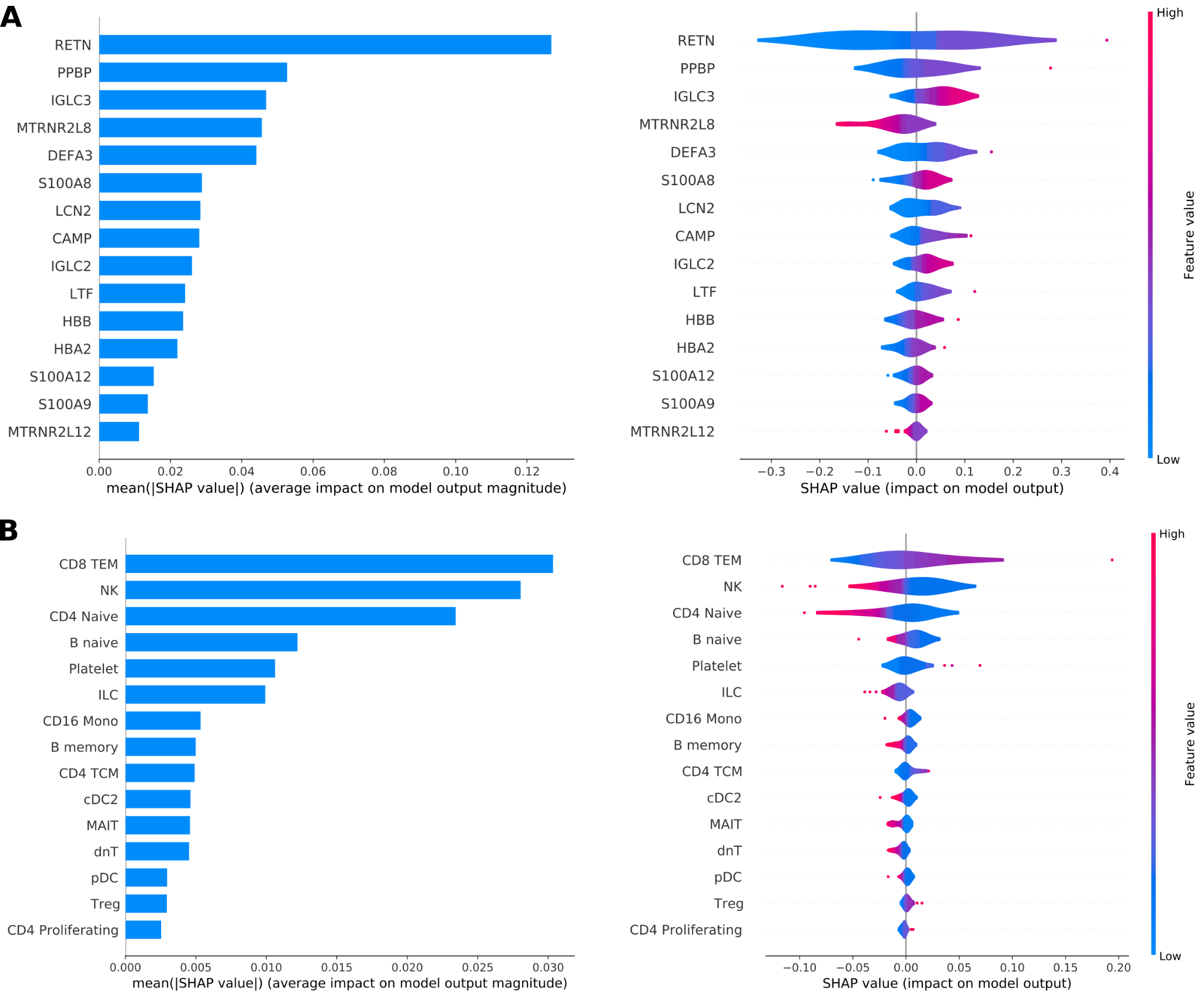

Figure S2: Feature importance based on SHapley Additive exPlanation (SHAP) values evaluated on the Stanford dataset. Bar plots represent the average across the 30 samplings of the mean absolute SHAP values to illustrate global feature importance (left). The violin plots show the direction of the relationship between each feature and the prediction outcome (right). The color in the violin plot represents the average feature value at that position. Features with many instances in red with SHAP values greater than 0 contribute positively to the prediction, while those with many blue instances decrease the prediction. (Related to figure 4)

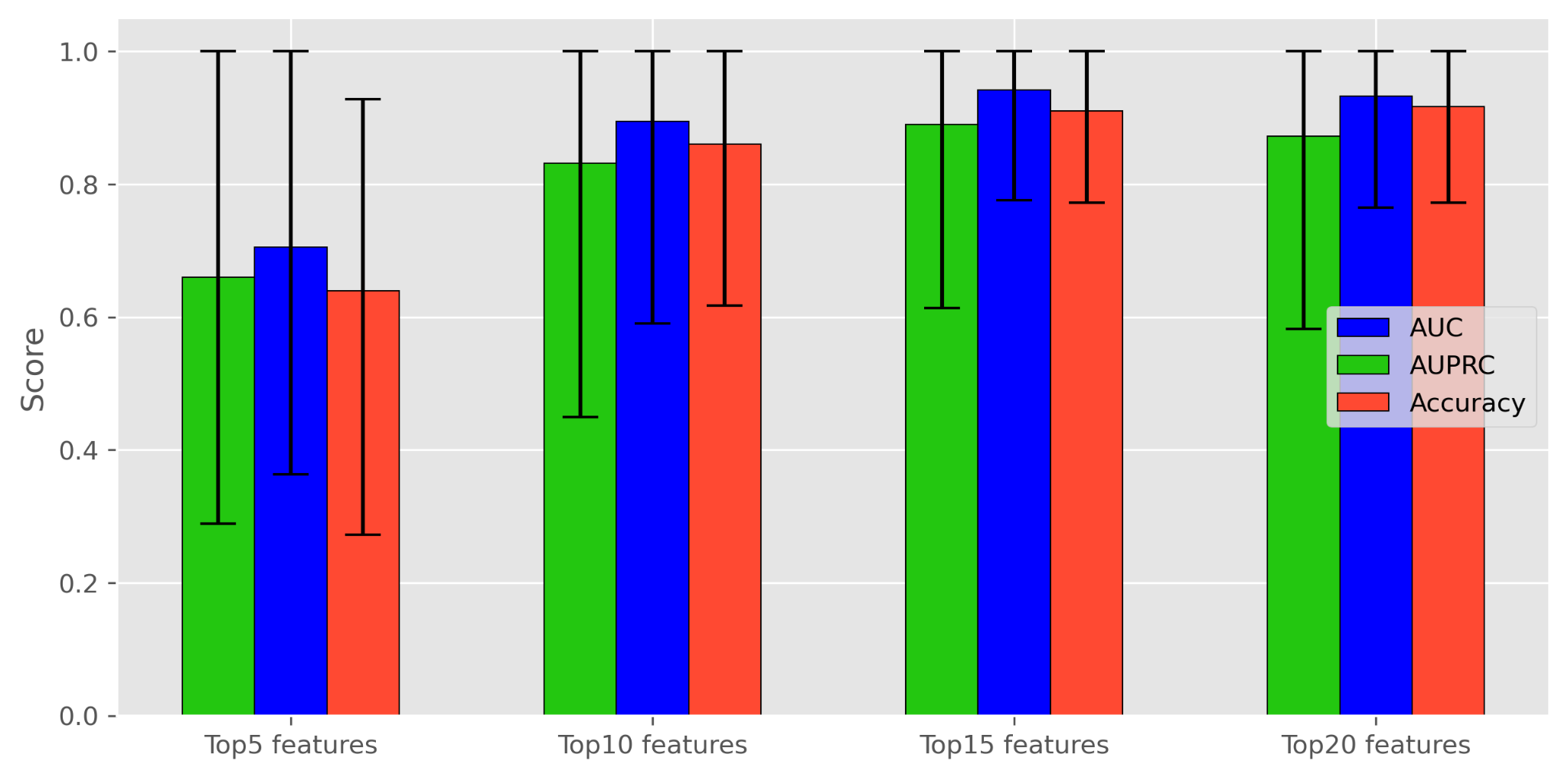

Figure S3: Gene-selection results from the MLP- joint model on 30 random samplings of the validation set. barplots show the mean AUC, AUPRC and Accuracy values and the error bars represent the Confidence Interval of 95% .
